# Histopathological features of hepatocellular carcinoma in patients with hepatitis B and hepatitis D viruses infection: A single-institution study from Mongolia

**DOI:** 10.1101/2024.10.18.24315628

**Authors:** Orgil Jargalsaikhan, Wenhua Shao, Mayuko Ichimura-Shimizu, Soichiro Ishimaru, Takaaki Koma, Masako Nomaguchi, Battogtokh Chimeddorj, Khongorzul Batchuluun, Anujin Tseveenjav, Battur Magvan, Sayamaa Lhagvadorj, Adilsaikhan Mendjargal, Lhagvadulam Ganbaatar, Minoru Irahara, Masashi Akaike, Damdindorj Boldbaatar, Koichi Tsuneyama

## Abstract

Viral hepatitis, particularly hepatitis B (HBV) and hepatitis C (HCV), is highly prevalent in Mongolia. Moreover, Mongolia has the highest prevalence of hepatitis delta virus (HDV) globally, with over 60% of HBV-infected individuals also co-infected with HDV. Since HBV/HDV infections accelerate liver disease progression more compared to HBV infection alone, urgent national health measures are required. This study presents a clinicopathological analysis of 49 hepatocellular carcinoma cases surgically resected at the Mongolia-Japan Hospital of the Mongolian National University of Medical Sciences. HBV infection was found in 27 (55.1%) cases of all HCC cases. Immunohistochemical staining of the liver revealed that 14 (28.6%) among the HBV infected tissues were HDV antigen-positive in the HCC cases. HDV-positive cases exhibited significantly higher inflammatory activity compared to HDV-negative cases, with lymphocytic infiltrates predominantly composed of CD4-positive cells. Furthermore, HDV-positive cells were spatially distinct from HBs antigen-positive cells, suggesting that HDV-infected cells may interfere with HBV replication. No significant differences in fibrosis or in tumor characteristics were observed between the HDV-positive and -negative cases.

Early diagnosis of HBV/HDV infections is essential for appropriate treatment and to prevent further domestic transmission of the virus. However, routine testing for HDV infection is rarely conducted in Mongolia. Since HDV-positive cells are morphologically indistinguishable from surrounding HDV-negative cells, routine histopathological analysis may not be sufficient to detect HDV infection. Based on this clinicopathological study, CD4 and CD8 immunostaining can be considered as an adjunctive diagnostic tool in cases with significant lymphocytic infiltration and hepatocellular damage. Additionally, HDV screening using blood and tissue samples may be recommended to ensure accurate diagnosis.

## Introduction

Liver disease in Mongolia presents a significant public health challenge, with the incidence rates of hepatocellular carcinoma (HCC) and cirrhosis the highest globally. A key factor behind this fact is the high prevalence of hepatitis B virus (HBV) and hepatitis D virus (HDV) co-infection among HBV carriers, which accelerates the progression of liver disease and increases the risk of hepatocellular carcinoma (HCC) (1-3). In Mongolia, more than 60% of HBV patients are also infected with HDV, a stark contrast to the global HBV/HDV infections rate of approximately 13% (4).

The HBV vaccine, introduced in the 1980s, remains the most effective preventive measure against HBV. The World Health Organization (WHO) recommends administering the first dose within 24 hours of birth, followed by subsequent doses according to national immunization schedules (5). This approach has significantly lowered HBV infection rates worldwide, particularly in high-risk regions such as Asia and Africa. For example, national vaccination campaigns in China and Taiwan have significantly reduced HBV transmission among younger populations (6,7). Similarly, Mongolia has adopted HBV vaccination programs, leading to a decrease in infection rates among younger demographics. However, challenges persist, especially in rural areas where knowledge about hepatitis is limited, leading to continued intra-family transmission from untreated carriers (8-10).

In contrast to HBV, no vaccine is currently available for hepatitis C virus (HCV). As HCV is primarily transmitted via blood, stringent infection control measures are crucial, particularly in medical settings. These include the use of disposable syringes, proper sterilization protocols, and rigorous testing of blood derivatives. Thorough reduction initiatives, such as providing sanitary needles to intravenous drug users, have proven effective in reducing HCV transmission in many countries, (11). Furthermore, ensuring regular screening, particularly in high-risk groups such as injecting drug users and those who have received blood transfusions, is critical for early detection and intervention.

To mitigate the impact of liver disease, Mongolia government initiatives such as the “Healthy Liver” campaign aim to raise public awareness and improve access to antiviral treatments (12). To assess the impact of these efforts and to better understand the epidemiological and clinical landscape of liver disease in the country, further investigation is needed.

This study examines 49 recently resected cases of HCC from the Mongolia–Japan Hospital of Mongolian National University of Medical Sciences (MNUMS) in Ulaanbaatar. Through clinical-pathological assessment and immunohistochemical analysis for HDV antigens, we aim to elucidate the morphological features of HBV/HDV infections and provide insights into the current state of HCC in Mongolia.

## Materials and Methods

### Selection of Cases

Seventy-two cases of HCC that underwent surgical resection were extracted from medical records of the Mongolia–Japan Hospital of MNUMS in Ulaanbaatar between August 2020 and July 2024. All cases were initially assessed by Mongolian pathologists (S.L.), and 22 cases, including cholangiocarcinoma and hemangioma, were excluded. One case of squamous cell carcinoma was excluded after being reevaluated by Japanese liver pathologist (K.T.). A total of 49 cases of HCC were analyzed (age: 65 [IQR 55, 69]; sex: male 27, female 22). For each case, clinical data including gender, age, liver function, blood biochemistry, and viral infection status, were collected. In addition, we analyzed three cases of serological HBV/HDV infections which were provided from National Pathology Center of Mongolia. Ethical approval for this study, involving clinical specimens, was obtained from the Medical Research Ethical Review Board at the Ministry of Health, Mongolia (#24/066, July 9, 2024).

### Histological Analysis

Histopathological evaluation was performed on hematoxylin and eosin (HE)-stained slides, selected from both cancerous and non-cancerous liver tissue (background liver). A consensus evaluation of liver pathology was performed by three expert liver pathologists (J.O., S.W., and K.T.,). For the assessment of tumor histology, activity of inflammation was graded on a 4-point scale (A0: no activity, A1: mild activity, A2: moderate activity and A3: severe activity), and fibrosis stage was assessed on a 5-point scale (F0: no fibrosis, F1: portal fibrosis without septa, F2: portal fibrosis with few septa, F3: numerous septa without cirrhosis and F4: cirrhosis) based on METAVIR score (13). If any notable variability in activity or fibrosis between different regions of the liver was detected, a mixed score (e.g., A1-A2) was assigned, and the median value was used for statistical purposes.

HCC characteristics were also evaluated in tumor tissues, focusing on the presence of clear cells, fat deposition, significant fibrosis, and vascular invasion. Each of these features was scored on a binary scale (presence or absence). The degree of lymphocytic infiltration within and around the tumor (intertumoral and peritumoral) was classified as either prominent or not prominent.

### Immunohistochemistry

Antibodies targeting hepatitis B surface antigen (HBsAg) and hepatitis D virus (HDV) antigens (both large and small delta antigens) were used. After the deparaffinization of the specimens, antigen retrieval was performed with microwave heating. After application of primary antibodies for mouse monoclonal anti-HBs (5C3, GeneTex, USA) and for rabbit monoclonal-anti HDV (large and small delta antigen, Abcam, UK), secondary antibodies polymer against mouse/rabbit IgG with peroxidase (Envision-PO for mouse or rabbit, Agilent, USA) were applied. Diaminobenzidine (DAB) reaction (Abcam) was done for the color of peroxidase. Counterstain was performed by hematoxylin.

Five cases with HDV antigen positive showing prominent lymphocytic infiltration were selected for double immunostaining using mouse monoclonal anti-CD4 antibody (NCL-CD4-1F6, Leica Biosystems, UK) and mouse monoclonal anti-CD8 antibody (Leica Biosystems) to assess the immune response within the liver tissue. The primary antibody (CD4) and secondary antibody (Envision-PO) were applied using the standard enzyme-antibody method, with CD4-positive cells labeled in brown using DAB as the substrate. The specimen was then immersed in PBS at a temperature of 95°C or higher for 15 minutes to inactivate the primary and secondary antibodies which were initially applied. CD8 antibodies and Envision-PO were subsequently applied, and CD8-positive cells were labeled in blue using the chromogenic substrate PermaBlue/HRP (DBS Diagnostic Biosystems, Netherlands).

### Statistical Analysis

All data are expressed as median (interquartile range), or frequency (percentages) as applicable. Histopathological features of the liver with HDV infection were compared between the two groups of positive and negative HDV antigens. One HCV co-infection case was included respectively in HDV positive and negative cases. Mann-Whitney U test was used to compare the continuous variables between the two groups. The chi-square test and Fisher’s exact test were used for evaluating any associations between categorical variables. A p-value of less than 0.05 was considered as statistically significant. All tests were conducted using IBM SPSS 29.0 (IBM, Armonk, NY).

## Results

### Clinical and Pathological Characteristics of HCC Cases

Comprehensive clinical data were available for most cases from the Mongolia–Japan Hospital of MNUMS, although serological data, particularly regarding HDV status, had defects in some cases (Supplemental Table 1). Therefore, HDV infection was defined as positive for HDV antigen (Ag) by immunohistochemical assessment instead of by serological diagnosis in the present study. The HBV infection was judged based on either serological diagnosis or immunohistochemical evaluation. The HBV infection was found in 27 cases (55.1%) of all HCC cases (Fig. 1). Among these HCC cases, 14 (28.6%) cases were HDV Ag-positive and one of these cases was co-infected with HCV. This suggests that HDV is involved in 51% of HBV-related HCC. The etiology of the 12 cases was unknown, as the viral status could not be evaluated. None of the cases exhibited signs of autoimmune hepatitis or primary biliary cholangitis. Although mild-moderate steatosis was observed in 9 cases, there was no histological evidence of steatohepatitis such as ballooning or perivenular-pericellular fibrosis. Hence, it was presumed that most HCC cases were mainly related to viral hepatitis.

**Figure 1.**
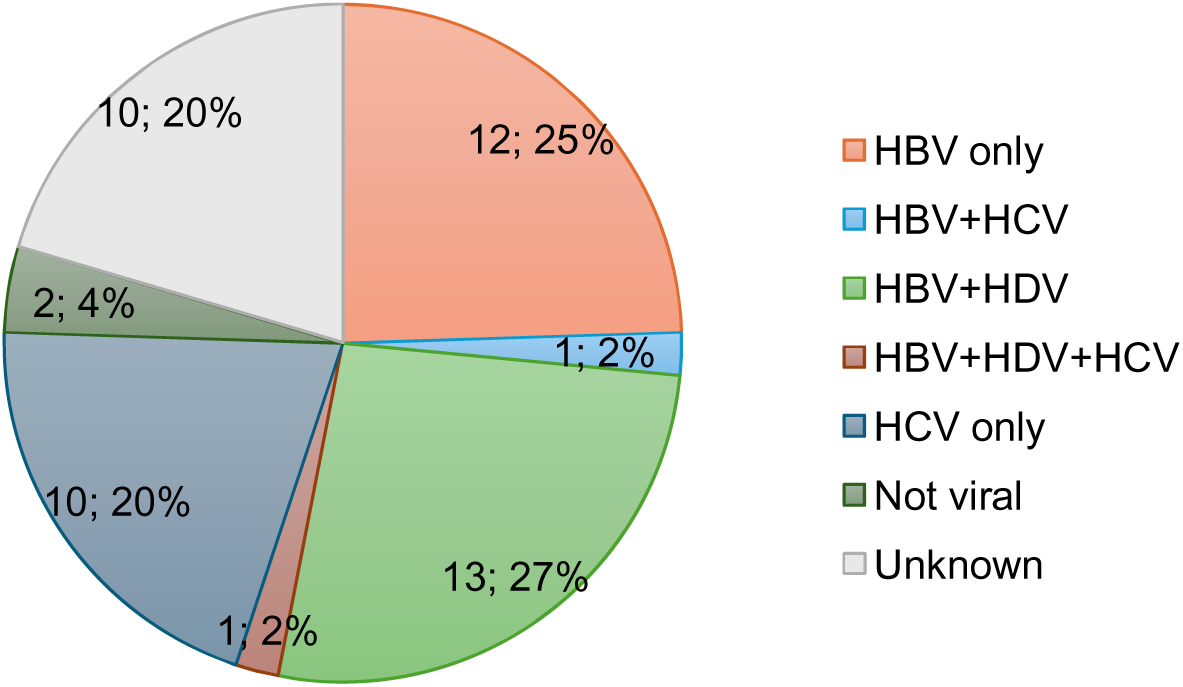
The proportion of viral hepatitis in HCC patients. The proportion of viral hepatitis based on liver histology or serology among the patients who had surgery for HCC. The numbers in the pie chart indicate the number of cases.

### Pathological Features of Background Liver Tissue in HDV-Positive Cases

The histopathological characteristics of the HCC cases are summarized in Supplemental Table 2. Figure 2A shows typical histological images of the tumor and background liver in HDV-positive cases. In the background liver, significant lymphoid infiltration was observed in fibrous septa, indicating severe interface hepatitis (Fig. 2B). Hepatocytes were damaged by infiltrating lymphocytes. Fibrous extension occurred from portal area in various degrees. In the HBV-related HCC, inflammatory activity in the background liver was notably elevated in HDV-positive cases compared to HDV-negative cases (Fig. 2C, P=0.043), although no significant difference in fibrosis severity was seen between the two groups (Fig. 2D).

**Figure 2.**
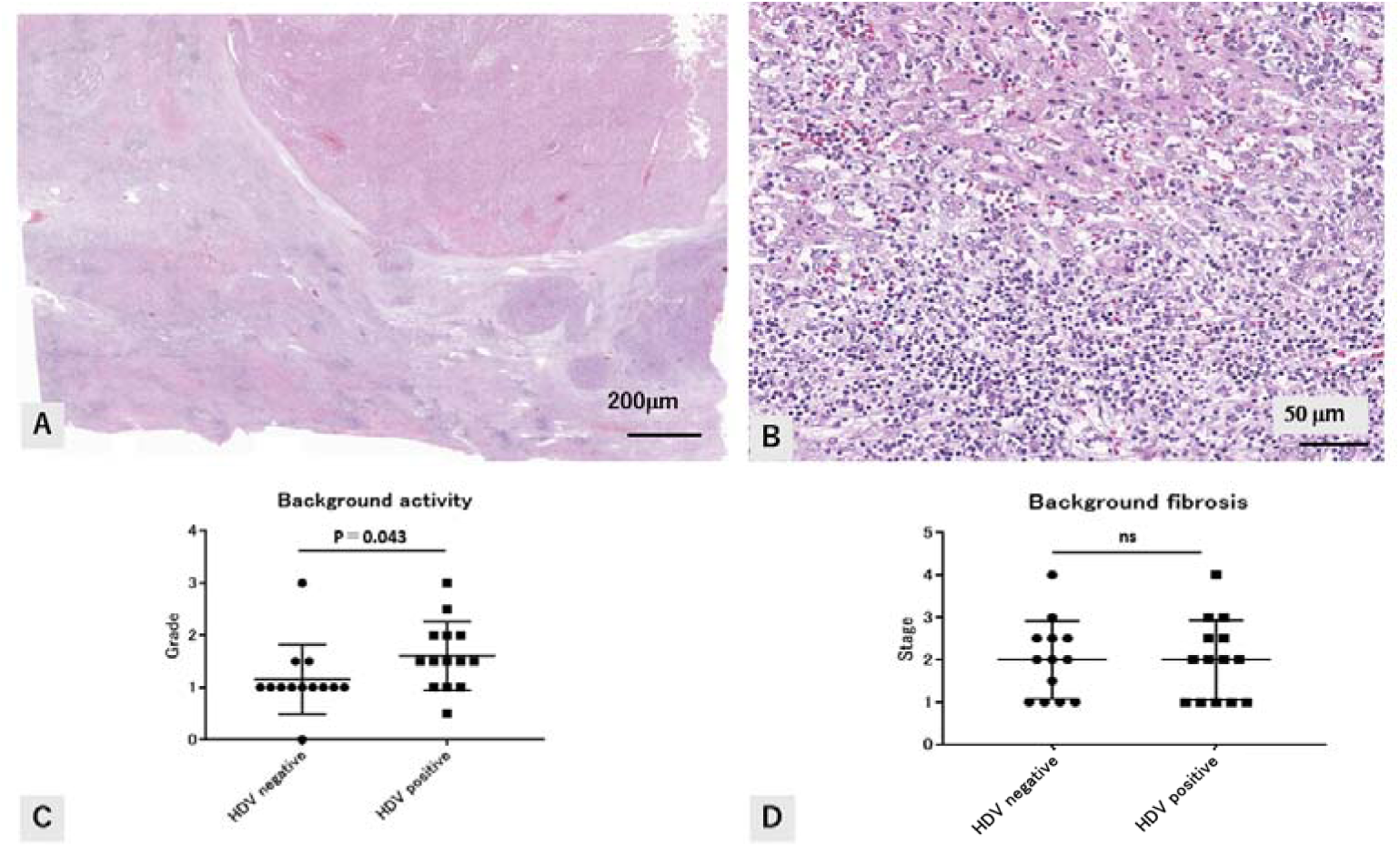
Pathological Features of Background Liver Tissue in HDV-Positive Cases. A, B: Representative images of HE staining of HDV-positive cases. A: Low-power view showing both the tumor and surrounding (background) liver tissue. Small regenerative nodules are visible around the primary encapsulated nodule. The surrounding liver tissue demonstrates significant inflammatory cell infiltration, accompanied by moderate fibrosis. (Scale bar: 200 μm) B: Higher magnification of the background liver tissue. A marked infiltration of lymphocytes is present, particularly at the interface, with clear evidence of hepatocyte damage due to the immune response. (Scale bar: 50 μm) C, D: Comparative analysis of inflammation and fibrosis scores in the background liver of HDV-positive and -negative cases. Inflammatory activity was significantly elevated in HDV-positive cases (P = 0.043).

Immunohistochemical analysis demonstrated a distinct segregation between hepatitis B surface antigen (HBsAg)-positive and HDV antigen-positive regions. HBsAg-positive cells were sparse in areas with diffused HDV positive regions, and conversely, HDV-positive cells were rarely present in regions with clustered HBsAg-positive cells (Fig. 3A-D). Morphologically, no cytoplasmic inclusions or other distinguishing features were observed by hematoxylin and eosin (HE) staining in areas containing HDV-positive cells. Notably, there was significant lymphocytic infiltration, particularly in pronounced interface hepatitis areas surrounding HDV-positive hepatocytes in the background liver (Fig. 4A). These infiltrating lymphocytes were predominantly CD4-positive T cells, with fewer CD8-positive cells (Fig. 4B, C). In contrast, an equal distribution of CD4-positive and CD8-positive cells was observed within tumors with no HDV-positive cells. HDV antigen expression was confined to hepatocytes and negative in interlobular bile ducts, except in one case where HDV antigen was detected in the nuclei of proliferating bile ductules (Fig. 5A, B).

**Figure 3.**
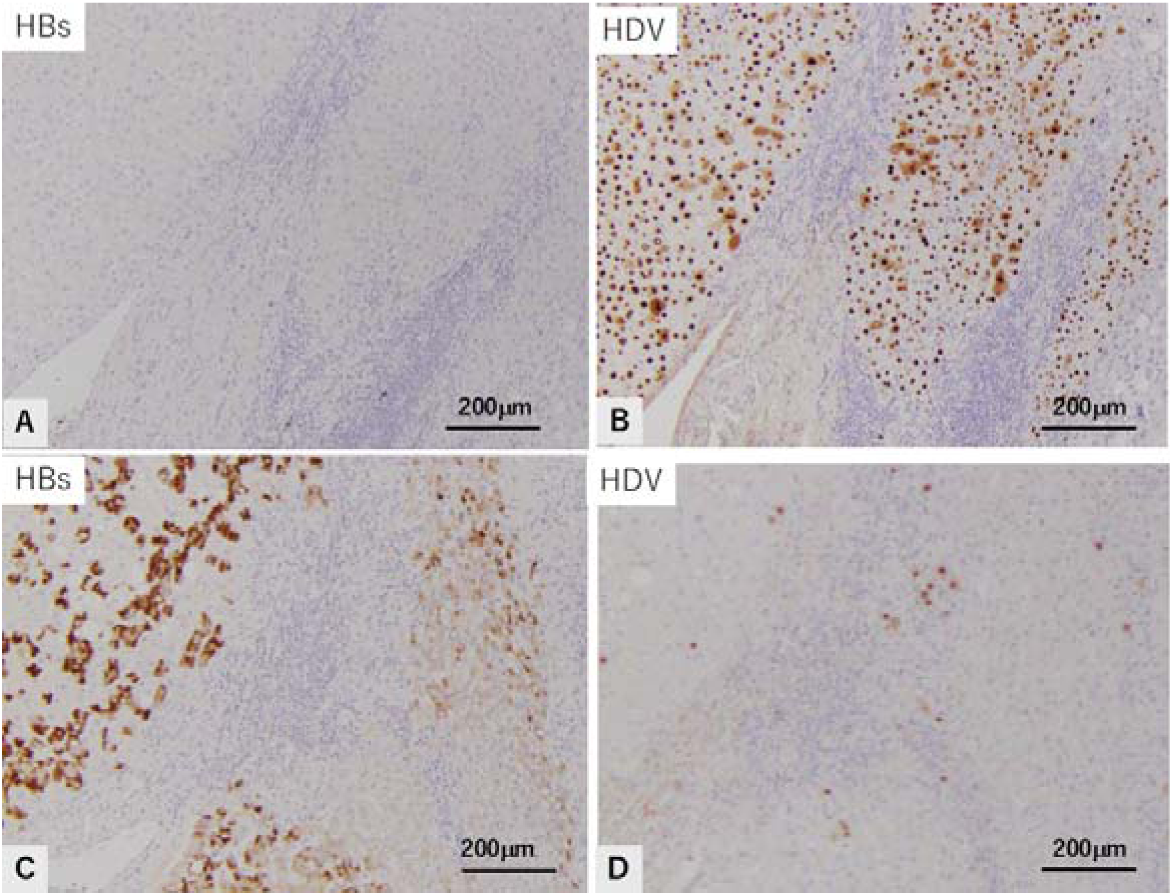
Immunohistochemical staining images for HBsAg and HDV on serial sections. A, C: HBsAg immunostaining; B, D: HDV immunostaining. A and B represent the same case, as do C and D. In A and B, only a few HBsAg-positive cells are present, while a large number of HDV-positive cells are observed. In contrast, C and D show clusters of HBsAg-positive cells, but only a few HDV-positive cells. Notably, there is minimal overlap between HBsAg-positive and HDV-positive cells. Scale bar: 200 μm

**Figure 4.**
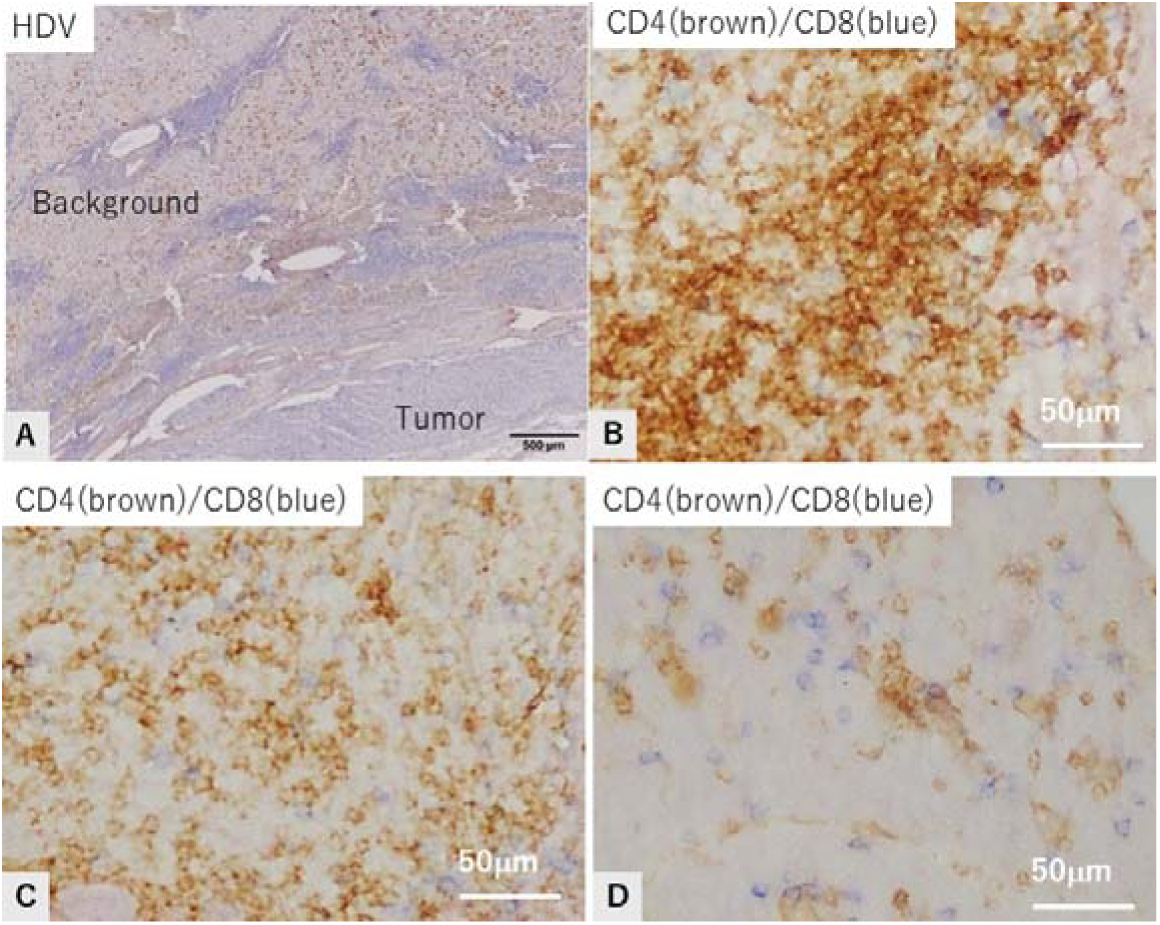
Lymphocytic profile surrounding HDV-positive hepatocytes. A: HDV immunostaining in the background liver shows widespread HDV-positive hepatocytes. Significant lymphocytic infiltration is observed, particularly at the interface hepatitis regions in the portal areas and within the fibrous septa. In contrast, HDV-positive cells are nearly absent within the tumor. B-D: Double immunostaining of lymphocytes (CD4 in brown, CD8 in blue) infiltrating the liver. B, C: In the background liver, the majority of infiltrating lymphocytes are CD4-positive, with only a few CD8-positive cells. D: In the tumor, the lymphocytic infiltration shows a more balanced ratio of CD4-positive and CD8-positive cells. Scale bar for A: 500 μm, B-D: 50 μm.

**Figure 5.**
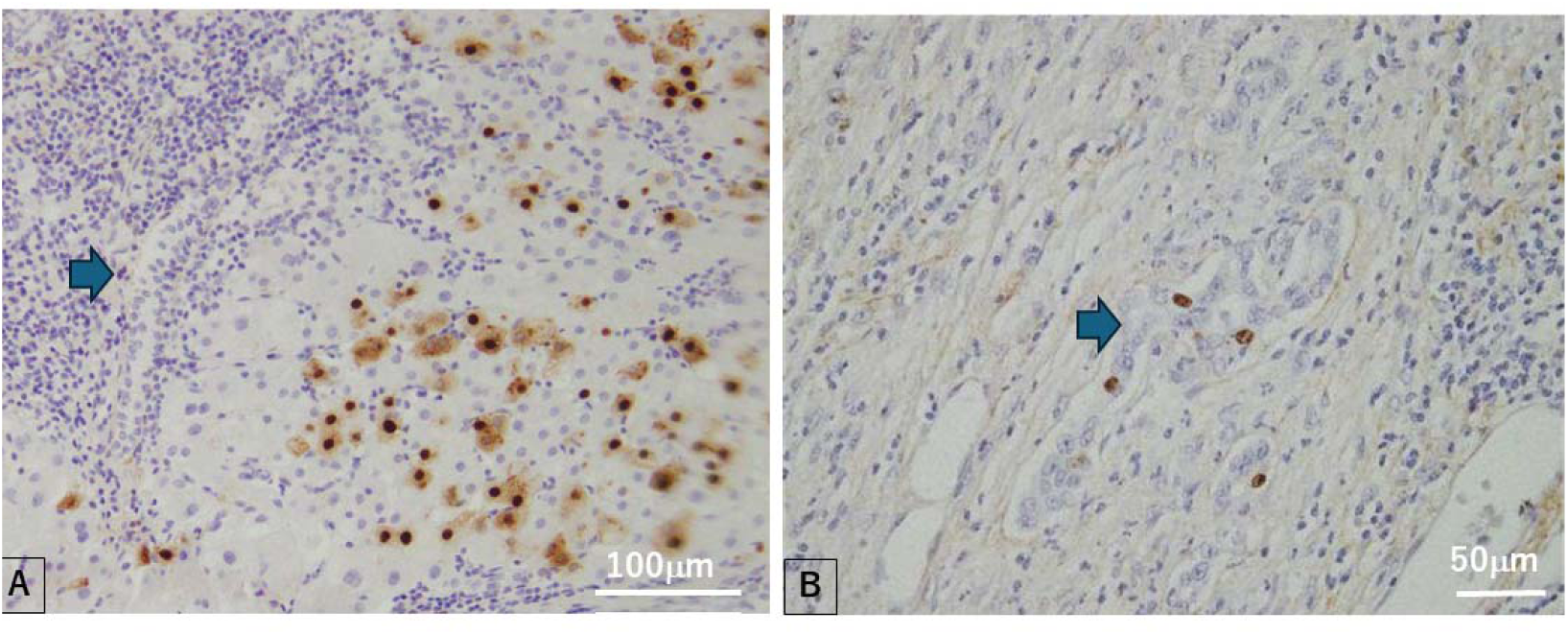
Immunohistochemical detection of HDV antigen in the biliary system. A: HDV-positive cells were confined to hepatocytes, with no HDV antigen expression observed in the interlobular bile ducts (arrow). B: In one case, HDV antigen was detected in the nuclei of proliferating bile ductules (arrow) located in the interface region with marked inflammation. Scale bar: A, 100 μm; B, 50 μm

### Pathological Features of Hepatocellular Carcinoma in HDV-Positive Cases

HCCs in this study were predominantly presented as simple nodular types, with a frequent nodule-in-nodule growth pattern. Tumors typically contained a mix of well-differentiated and moderately differentiated components, with poorly differentiated or undifferentiated elements rarely observed (Supplemental Table 2). No significant differences in tumor characteristics such as clear cell type, fat deposition, intertumoral fibrosis, lymphocytic infiltration (both intertumoral and peritumoral cuffing), and vascular invasion were seen between HDV-positive and HDV-negative cases. However, there was a trend towards increased fat deposition within the tumor tissue in HDV-positive cases, although this difference did not reach statistical significance (P = 0.066, Table 1). While most HCCs contained only a few HDV antigen-positive cells (Fig. 6C), in two of the cases, the number of HDV-positive cells was higher within the tumor tissue compared to the surrounding liver tissue (Fig. 6D).

**Figure 6.**
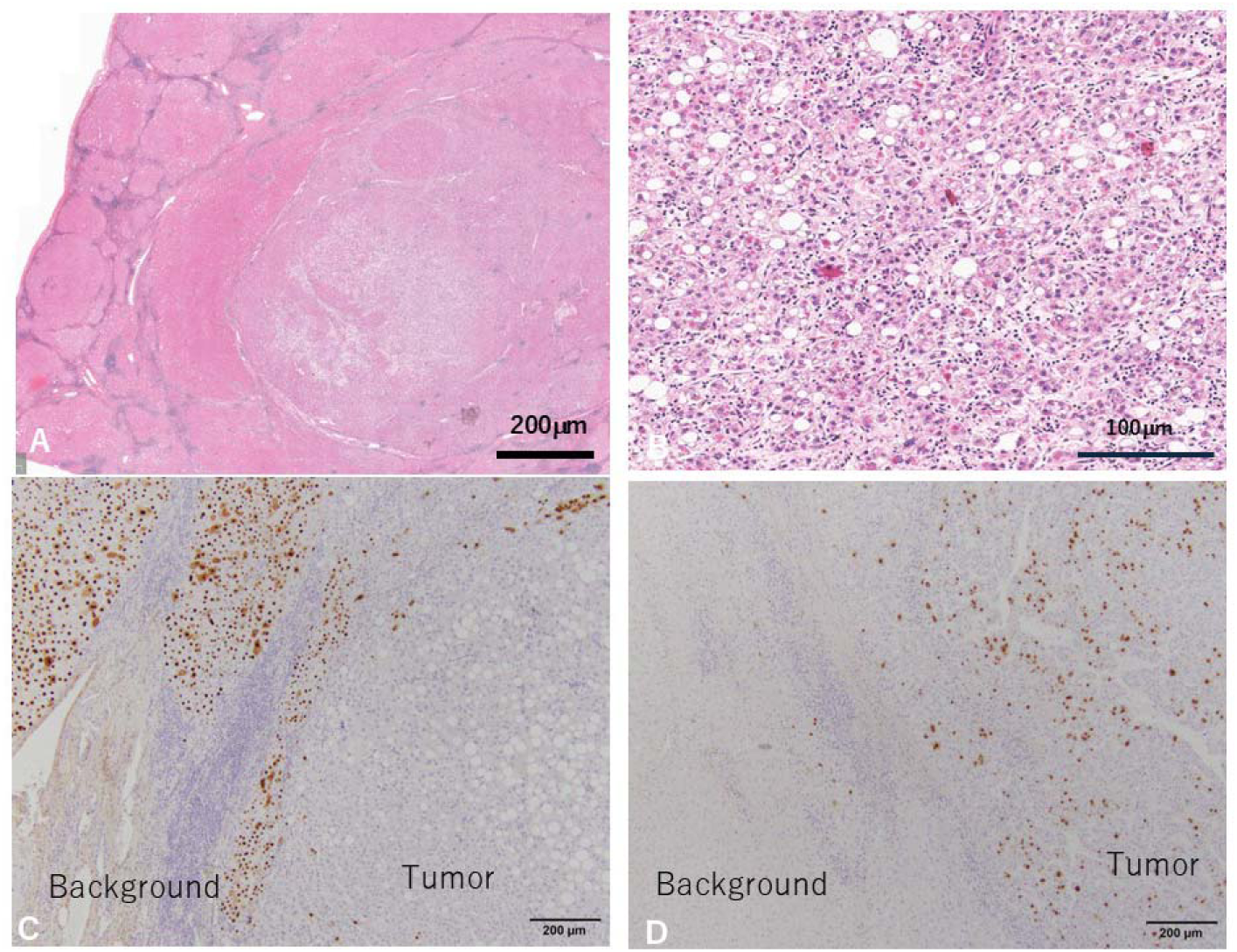
Characteristics of liver tumors in HDV-positive cases. A, B: Representative images of liver tumors (A: low magnification, B: high magnification of the tumor center). C, D: HDV immunohistochemistry. Most hepatocellular carcinomas (HCCs) showed only a small number of HDV-positive cells (C). However, in two cases, the number of HDV-positive cells within the tumor tissue exceeded that in the surrounding liver tissue (D). Scale bar: A,C and D, 200 μm; B, 100 μm.

**Table 1.** Observed number of cases with lipid droplet in the liver tumor.

|  | Lipid droplet in the tumor |  | Total |
| --- | --- | --- | --- |
|  | Absent | Present |  |
| HDV-positive | 11 | 2 | 13 |
| HDV-negative | 7 | 7 | 14 |
| Total | 18 | 9 | 27 |

P=0.057 by Fisher's exact test

## Discussion

HDV, a satellite virus reliant on the HBV, causes the most severe form of chronic hepatitis and is associated with significant morbidity and mortality (14). The clinical spectrum of HDV infection ranges from asymptomatic to acute liver failure, leading to chronic hepatitis, with rapid progression to cirrhosis, liver failure, or HCC, exceeding the extent of severity typically observed in HBV infection alone. Although HBV/HDV infections can sometimes resolve spontaneously, most superinfection carriers of HBV with HDV develops chronic HDV infection (15).

The assembly and release of virus particles, as well as HDV entry into hepatocytes, depend on the presence of the HBV S protein. However, HBV is not required for HDV replication, as HDV can replicate in the nucleus by interacting with host proteins (14). Chronic hepatitis D is characterized by a severe and progressive disease course, with cirrhosis developing in up to 80% of the cases, often culminating in liver failure or HCC (16). HDV exacerbates HBV-associated liver pathology, accelerating processes such as inflammation, fibrosis, and cirrhosis. Recent research has indicated that HDV enhances oxidative stress and endoplasmic reticulum (ER) stress caused by HBV (17).

In the present study, viral hepatitis accounts for most HCC cases in Mongolia, with HDV implicated in approximately one-third. Histological analysis revealed moderately-to severely lymphocytic infiltration around damaged HDV-positive hepatocytes, suggesting that immune responses to HDV antigens may contribute significantly to the severity of HDV-associated liver disease (18). Notably, HDV-positive cases exhibited increased lymphocytic infiltration, particularly involving CD4^+^ T cells rather than CD8^+^ T cells. This finding is consistent with previous reports describing predominant infiltration of cytotoxic CD4^+^ T cells expressing perforin around the HDV-infected cells (17). The complex interplay between the immune response and viral evolution in chronic HDV infection is notable. While the immune response to HDV is heavily mediated by CD8^+^ T cells, the data also suggests a significant role for CD4^+^ T cells in the context of persistent infection. This impaired response may be a consequence of viral mutations that allow HDV to escape CD8^+^ T cell recognition, as discussed in studies where HDV variants evolve to avoid detection by common HLA class I molecules. These immune escape mechanisms reduce the effectiveness of CD8^+^ T cell responses, potentially leading to an increased involvement of CD4^+^ T cells in the immune response. A predominant infiltration of cytotoxic CD4^+^ T cells expressing perforin in the liver could compensate for the diminished CD8^+^ T cell function and target HDV-infected cells. The shift toward CD4^+^ T cell involvement may reflect an adaptive immune strategy as the virus evades typical CD8^+^ T cell-mediated clearance. Understanding these immune dynamics is crucial for developing new therapeutic approaches that aim to restore effective CD8^+^ T cell responses and to harness the role of CD4^+^ T cells in controlling chronic HDV infection (19.20). Although this study did not observe significant differences in fibrosis between HDV-positive and HDV-negative cases, other studies have documented accelerated fibrosis in HDV infection (8.9). The presence of active inflammation and fibrosis, particularly in interface hepatitis dominated by CD4^+^ T cells, may indicate HBV/HDV infections.

Interestingly, in HDV-positive cases, HBsAg-positive cells and HDV antigen-positive cells were spatially distinct, suggesting that HDV may interfere with HBV replication. Previous research by Chiba et al. demonstrated that HDV-infected hepatocytes activate sustained interferon (IFN) responses, leading to significant inhibition of HBV replication (21). This observation holds important implications for therapeutic strategies that targets both HBV and HDV infections.

Regarding hepatocarcinogenesis, it has been proposed that the development mechanisms differs by which HBV and HDV contribute to tumor. (15). In most cases, the number of HBs-positive and HDV-positive cancer cells was significantly lower than in the surrounding liver tissue, suggesting that both viruses may promote tumor initiation rather than promoting progression. However, in two HDV-positive cases, the number of HDV-positive cancer cells was significantly higher than in the surrounding background hepatocytes, indicating the possibility of HDV replication within the tumor cells. Future studies involving genetic analyses using microdissection systems are essential to further elucidate the virological status in cancer cells in these cases.

Another unresolved issue is the nuclear expression of HDV antigen in proliferating bile ductulus. In this study, no other HDV expression was observed in the biliary system, and no cases including differentiation of cholangiocarcinoma were identified. Proliferating bile ductulus are considered to have originated either from precursor/stem cells or from metaplasia of mature hepatocytes during periods of active interface hepatitis. It is plausible that HDV-positive proliferating bile ductulus represent metaplasia from HDV-infected hepatocytes.

The backgrounds of the remarkably high prevalence of HBV/HDV infections in the Mongolian population remains a mystery. Identifying the factors contributing to this phenomenon is a critical priority for controlling HDV in Mongolia. There is a big difference between the medical environment in rural and urban areas in Mongolia, and there still remains parts of rural areas where education on infectious diseases is insufficient. Access to treatment for hepatitis in Mongolia remains limited due to a lack of screening, clinical services, and the high cost of antiviral drugs. While the price of medications for hepatitis B and C has dropped by nearly 90%, they still remain out of reach for the majority of Mongolians (22). In addition, the recent discovery of HDV-like viruses in various animal species, including rodents and snakes, challenges the previous understanding of HBV because the sole helper virus for HDV replication may exist. Given the common consumption of wildlife, such as marmots, in Mongolia, animal vectors may play a role in HDV transmission (23). Comprehensive epidemiological studies are required to investigate the potential involvement of animal reservoirs in HDV transmission.

## Data Availability

All data produced in the present work are contained in the manuscript

## Author Approvals

The authors certify that the manuscript has not been submitted elsewhere for publication. All authors have seen and agreed to submit the manuscript.

## Acknowledgements

We are grateful to Japan International Cooperation Agency (Japan) for the support via Project for Strengthening of Hospital Management and Education for Medical Staff at the Mongolia-Japan Hospital. This work was supported by funding from the Research Clusters program of Tokushima University (T.K.), the Takeda Science Foundation (T.K.) and the Heiwa Nakajima Foundation (T.K.).

