## Supplemental Table 2 for "Histopathological features of hepatocellular carcinoma in patients with hepatitis B and hepatitis D viruses infection: A single-institution study from Mongolia": SupplementalTable2.docx

| Supplemental Table 2 Individual Histopathological features of HCC patients | | | | | |  |  |  |  |  |  |  |  |
| --- | --- | --- | --- | --- | --- | --- | --- | --- | --- | --- | --- | --- | --- |
| Viral infection |  | Background liver tissue | | |  | Tumor tissue | | | | | | | |
|  | No. | Inflammatory activity | Fibrosis | Remarks |  | Differentiation grade | Tumor clear cells | Tumor fat | Intratumor fibrosis | Vascular invasion | Intratumor lymhocytes | Peritumor-cuffing lymphocytes | Remarks |
| HBV | 3 | 1 | 2 |  |  | mod | - | - | - | - | - | - |  |
|  | 6 | 1 | 2.5 |  |  | wel-mod | + | - | - | - | + | - |  |
|  | 9 | 1 | 2.5 |  |  | wel | - | + | - | - | - | - |  |
|  | 10 | 1 | 1 | mild steatosis |  | wel | + | + | - | - | - | - |  |
|  | 15 | 1 | 1 |  |  | mod | + | - | - | + | + | - |  |
|  | 17 | 1 | 2 |  |  | wel-mod | - | - | - | - | - | + |  |
|  | 20 | 0 | 1.5 | steatosis |  | mod | - | - | - | - | - | - |  |
|  | 30 | 1.5 | 1 |  |  | mod | - | - | - | + | - | + |  |
|  | 31 | 3 | 2.5 |  |  | mod | - | - | - | + | + | - |  |
|  | 33 | 1 | 3 |  |  | mod | - | - | - | - | - | - |  |
|  | 41 | 1 | 1 | mild steatosis |  | mod | - | - | - | - | - | + |  |
| HCV | 8 | 1 | 1 |  |  | mod | - | - | + | + | - | + |  |
|  | 21 | 1 | 0 |  |  | mod | + | - | - | - | + | - |  |
|  | 22 | 1 | 1.5 |  |  | mod | + | - | - | + | - | - |  |
|  | 26 | 2 | 1.5 |  |  | mod-por | - | - | - | + | + | - |  |
|  | 27 | 1 | 1 |  |  | mod | + | - | - | - | + | - |  |
|  | 32 | 2.5 | 2.5 |  |  | mod | - | + | + | + | + | - |  |
|  | 35 | 1 | 2.5 | steatosis |  | mod | - | - | + | + | - | - |  |
|  | 37 | 1.5 | 4 | mild steatosis |  | mod | + | + | - | + | - | - |  |
|  | 38 | 0.5 | 0.5 | NRH |  | wel-mod | - | - | - | - | + | - |  |
|  | 48 | 1 | 1 |  |  | mod | - | - | - | - | - | - |  |
| HBV+HCV | 1 | 1 | 2 |  |  | wel-mod | - | - | - | + | + | - |  |
| HBV+HDV | 4 | 1.5 | 2.5 |  |  | wel | - | + | - | - | + | - |  |
|  | 7 | 1 | 1 |  |  | mod | - | + | - | - | - | - |  |
|  | 11 | 3 | 2 |  |  | mod | + | + | - | + | - | - |  |
|  | 12 | 1.5 | 2 |  |  | wel-mod | - | + | - | - | - | - | Necrosis |
|  | 23 | 0.5 | 1 |  |  | mod | - | - | - | - | - | - |  |
|  | 29 | 1.5 | 1 | mild steatosis |  | mod | - | + | - | - | - | + |  |
|  | 34 | 2 | 2.5 |  |  | wel-mod | - | + | - | - | + | - |  |
|  | 36 | 2.5 | 4 | mild steatosis |  | mod | + | - | - | - | + | - |  |
|  | 39 | 1 | 1 |  |  | mod | + | - | - | + | - | - |  |
|  | 42 | 1.5 | 1 |  |  | mod | + | - | - | + | - | - |  |
|  | 43 | 1 | 2 | mild steatosis |  | wel | - | + | - | - | + | - |  |
|  | 44 | 1.5 | 3 |  |  | mod | + | - | - | - | - | - |  |
|  | 45 | 2 | 2 |  |  | wel-mod | + | - | - | + | - | - |  |
|  | 49 | 1.5 | 4 |  |  | mod | + | - | - | - | - | - |  |
| HBV+HCV+HDV | 40 | 2 | 3 |  |  | mod | + | - | - | + | - | - |  |
| No niral | 18 | 1 | 0 |  |  | mod-por | - | - | - | + | - | - |  |
|  | 47 | 1 | 1 |  |  | mod | - | - | - | + | - | - |  |
| Unknown | 2 | 2.5 | 4 |  |  | mod | + | - | + | + | + | - |  |
|  | 5 | 1 | 1 |  |  | mod | + | - | + | + | - | - |  |
|  | 13 | 1 | 3.5 |  |  | wel-mod | + | - | - | - | + | - |  |
|  | 14 | 1.5 | 3 | mild steatosis |  | wel-mod | + | + | - | - | + | - |  |
|  | 16 | 1 | 0.5 |  |  | mod | + | + | - | + | + | - |  |
|  | 19 | 1.5 | 1 |  |  | mod | - | - | - | + | - | + |  |
|  | 24 | 1 | 3.5 |  |  | mod | - | - | - | - | - | - |  |
|  | 25 | ND | ND | No background |  | mod | + | + | + | + | - | + |  |
|  | 28 | 2 | 2.5 | steatosis |  | wel-mod | - | - | - | - | + | - |  |
|  | 46 | 1 | 1 |  |  | mod | - | - | - | - | + | - |  |
| The differentiation of liver tumors was classified into 3 grade: well-differentiated (wel), moderately-differentiated (mod), and poorly-differentiated (por).When two different grades of differentiation were observed within a tumor, the two grades were listed side by side. HBV, hepatitis B virus; HCV, hepatitis C; HDV, hepatitis D virus; NRH, nodular regenerative hyper-plasia | | | | | | | | | | | | | |
