## Supplemental Table 1 for "Histopathological features of hepatocellular carcinoma in patients with hepatitis B and hepatitis D viruses infection: A single-institution study from Mongolia": SupplementalTable1(rev).docx

| Supplemental Table 1 Individual Clinical data of HCC patients | | | | | | |  | |  |  | |  | | |  | |
| --- | --- | --- | --- | --- | --- | --- | --- | --- | --- | --- | --- | --- | --- | --- | --- | --- |
| No. | Blood biochemistry | | | |  | Viral infection | | | | | | |  | Immunohistochemistry | | |
|  | WBC (10^9^/L) | ALT (U/L) | AST (U/L) | Glucose (mmol/L) |  | HBV | | HDV | | | HCV | |  | HBs | | HDV |
| 1 | 7.12 | 44.37 | 42.88 | 4.88 |  | + | | ND | | | + | |  | ND | | ND |
| 2 | 16.32 | 128.9 | 308 | ND |  | ND | | ND | | | ND | |  | - | | - |
| 3 | ND | ND | ND | ND |  | ND | | ND | | | ND | |  | + | | - |
| 4 | 12.16 | 128.69 | 120.92 | ND |  | ND | | ND | | | ND | |  | + | | + |
| 5 | 6.96 | 71.1 | 41.73 | 5.19 |  | ND | | ND | | | ND | |  | - | | - |
| 6 | ND | ND | ND | ND |  | ND | | ND | | | ND | |  | + | | - |
| 7 | ND | ND | ND | ND |  | ND | | ND | | | ND | |  | + | | + |
| 8 | 5.67 | 12.28 | 18.44 | 5.22 |  | ND | | ND | | | + | |  | - | | - |
| 9 | ND | ND | ND | ND |  | ND | | ND | | | ND | |  | + | | - |
| 10 | ND | ND | ND | ND |  | ND | | ND | | | ND | |  | + | | - |
| 11 | 14.44 | 87.79 | 165 | ND |  | ND | | ND | | | ND | |  | + | | + |
| 12 | 23.88 | 64.47 | 330.37 | ND |  | + | | ND | | | - | |  | + | | + |
| 13 | 7.37 | 7.8 | 25 | ND |  | ND | | ND | | | ND | |  | - | | - |
| 14 | 4.49 | 1.56 | 28.4 | 6.25 |  | ND | | ND | | | ND | |  | ND | | ND |
| 15 | 3.72 | 40 | 19.89 | ND |  | ND | | ND | | | ND | |  | + | | - |
| 16 | 10.09 | 648 | 743 | ND |  | ND | | ND | | | ND | |  | ND | | ND |
| 17 | 3.8 | 18.62 | 15.95 | 5.57 |  | ND | | ND | | | ND | |  | + | | - |
| 18 | 23.3 | 48.16 | 98.79 | ND |  | - | | ND | | | - | |  | - | | - |
| 19 | ND | ND | ND | ND |  | ND | | ND | | | ND | |  | ND | | ND |
| 20 | 5.6 | 32.32 | 22.57 | 4.42 |  | ND | | ND | | | ND | |  | + | | - |
| 21 | 9.67 | 37.42 | 37.3 | ND |  | - | | ND | | | + | |  | ND | | - |
| 22 | 5.74 | 144.3 | 65.2 | ND |  | - | | ND | | | + | |  | - | | - |
| 23 | 4.89 | 507 | 231 | ND |  | + | | ND | | | - | |  | + | | + |
| 24 | 6.33 | 11.42 | 14.38 | 5.36 |  | ND | | ND | | | ND | |  | - | | - |
| 25 | 15.88 | 21 | 130 | ND |  | ND | | ND | | | ND | |  | - | | - |
| 26 | 11.62 | 173.71 | 217.42 | ND |  | - | | ND | | | + | |  | ND | | ND |
| 27 | 4.72 | 35.8 | 36.94 | ND |  | - | | ND | | | + | |  | - | | - |
| 28 | 10.64 | 21 | 11.16 | ND |  | - | | ND | | | ND | |  | - | | - |
| 29 | 5.44 | ND | ND | ND |  | + | | ND | | | ND | |  | + | | + |
| 30 | 10.09 | 648 | 743 | ND |  | ND | | ND | | | ND | |  | + | | - |
| 31 | 4.79 | 374 | 192 | 4.79 |  | + | | ND | | | - | |  | ND | | - |
| 32 | 10.68 | 52 | 61 | 5.67 |  | - | | ND | | | + | |  | - | | - |
| 33 | ND | ND | ND | ND |  | + | | ND | | | - | |  | - | | - |
| 34 | 3.97 | 74.91 | 64 | 4.98 |  | + | | + | | | - | |  | + | | + |
| 35 | 10.82 | 12.59 | 73.16 | - |  | - | | ND | | | + | |  | - | | - |
| 36 | 4.41 | 134 | 107.29 | 5.44 |  | + | | ND | | | - | |  | + | | + |
| 37 | 12.16 | 150 | 170 | 5.45 |  | - | | ND | | | + | |  | - | | - |
| 38 | 9.11 | 37.91 | 24.05 | 4.6 |  | - | | ND | | | + | |  | - | | - |
| 39 | 8.18 | 315 | 168.83 | ND |  | + | | ND | | | - | |  | + | | + |
| 40 | 6.64 | 186.47 | 172.28 | ND |  | + | | ND | | | + | |  | + | | + |
| 41 | 8.29 | 19.4 | 21 | ND |  | + | | ND | | | - | |  | - | | - |
| 42 | 9.02 | 46.34 | 46.24 | ND |  | ND | | ND | | | ND | |  | + | | + |
| 43 | 4.17 | 117.32 | 92.26 | 4.79 |  | + | | ND | | | - | |  | + | | + |
| 44 | 4.5 | 37.38 | 51.49 | ND |  | ND | | ND | | | ND | |  | + | | + |
| 45 | ND | ND | ND | ND |  | ND | | ND | | | ND | |  | + | | + |
| 46 | 9.07 | 33.94 | 32.33 | ND |  | ND | | ND | | | ND | |  | ND | | ND |
| 47 | 7.84 | 11.24 | 13.36 | ND |  | - | | ND | | | - | |  | ND | | ND |
| 48 | 6.39 | 139 | 210 | 7.4 |  | - | | ND | | | + | |  | - | | - |
| 49 | ND | ND | ND | ND |  | ND | | ND | | | ND | |  | + | | - |
| ALT, alanine aminotransferase; AST, aspartate aminotransferase; HBs, hepatitis B surface antigen; HBV, hepatitis B virus;  HCV, hepatitis C; HDV, hepatitis D virus; ND: Not Done; WBC, white blood cell | | | | | | | | | | | | | | | | |
